# Typhinder: Rapid, low-cost colorimetric detection of *Salmonella* Typhi bacteriophages for environmental surveillance

**DOI:** 10.1101/2025.01.13.25320463

**Authors:** Kesia Esther Da Silva, Tuya Yokoyama, Shiva Ram Naga, Mamata Maharjan, Paulo César Pereira dos Santos, Karla Fisher, Jean T. Coulibaly, Max Zhang Yang, Eric Jorge Nelson, Richelle C Charles, Katherine Shafer, Brad-Lot Igiraneza, Samaila Yusuf, Elisabeth Mulder, Kathleen Neuzil, Isaac I. Bogoch, Rajeev Shrestha, Dipesh Tamrakar, Jason R. Andrews

## Abstract

Typhoid fever remains a global public health challenge, especially in low- and middle-income countries where poor sanitation and limited access to clean water facilitate transmission. The lack of data on disease burden poses a significant barrier to adopting effective interventions such as vaccination programs. We developed a novel colorimetric assay for the detection of *S*. Typhi-specific bacteriophages (phages) in environmental water samples, providing an indirect indicator of *S.* Typhi contamination and insights into typhoid burden. We collected surface water samples from Brazil, Côte d’Ivoire, Nepal, and Niger, covering urban, peri-urban, and rural areas. We evaluated the colorimetric assay efficiency against agar overlay plaque assay. Isolated phages were tested against various bacteria to assess their host range. The colorimetric assay demonstrated high sensitivity (100% concordance with double agar overlay) with a detection limit of 28 plaque-forming units per milliliter (PFU/mL), and results were obtained in 5.5 hours. Phage detection rates were highest in densely populated areas with poor sanitation, particularly in Kathmandu, Nepal (96.6% positivity in river samples) and Abidjan, Côte d’Ivoire (35.1% positivity in drainage samples). The detection of *S*. Typhi phages in Côte d’Ivoire is particularly important, as the burden of typhoid in the region was previously undocumented. Phages were not detected in rural and drinking water sources. Host range analyses demonstrated that the isolated phages were specific to *S.* Typhi, with a small minority of phages (4/30) isolated also capable of infecting *S*. Paratyphi A. The novel colorimetric assay offers a rapid and sensitive method for detecting *S.* Typhi bacteriophages in environmental water. The scalability, low cost (∼$2.40 USD per sample), and minimal equipment requirements, suggest that this could be effective tool for typhoid surveillance in resource-limited settings.

**IMPORTANCE:** Typhoid fever, caused by *Salmonella* Typhi, remains a significant global health threat, particularly in low-resource settings with inadequate sanitation. Effective control measures, such as vaccines, require precise data on where typhoid is most prevalent, yet current surveillance methods are expensive and limited in scope. This study introduces a rapid, low-cost, and scalable colorimetric assay for detecting *S.* Typhi bacteriophages (viruses that infect bacteria) in environmental water samples. Unlike traditional methods, this test detects the presence of *S.* Typhi indirectly by identifying associated phages, offering a specific and sensitive approach for monitoring typhoid fever circulation. The assay was validated in diverse settings across four countries and demonstrated high accuracy and cost efficiency. By reducing reliance on expensive laboratory equipment and complex procedures, this tool makes typhoid surveillance more accessible, especially in low-income regions, helping prioritize vaccination campaigns and improve public health interventions.

## Introduction

Typhoid fever, caused by *Salmonella enterica* serovar Typhi (*Salmonella* Typhi), remains a major global health problem with an annual incidence of approximately 11 million cases and over 100,000 deaths worldwide (1). The burden is disproportionately high in low- and middle-income countries (LMICs) where access to clean water and sanitation is limited (2). Moreover, escalating antimicrobial resistance among *S*. Typhi strains poses serious challenges to effective treatment and may lead to higher mortality and greater strain on already overburdened healthcare systems (3). In response, the World Health Organization (WHO) recommends the introduction of typhoid conjugate vaccines (TCVs) in countries with a high-burden or disease and/or drug-resistant *S*. Typhi (4). However, a critical barrier to effective vaccine implementation is the scarcity of comprehensive data on typhoid burden in many endemic areas. Current estimates rely on short-term studies, often limited geographically and temporally (2,5). To overcome these limitations, there is growing recognition of the role of environmental surveillance as a complementary tool to identify high-risk areas and contribute crucial data for targeted interventions including vaccination (6,7).

Advancements in molecular techniques have enhanced the detection of *S*. Typhi DNA in environmental samples, although at considerable cost and requiring specialized laboratory infrastructure (8). Recently, there has been renewed interest in bacteriophages as a promising alternative for environmental monitoring of pathogenic bacteria like *S*. Typhi (9,10). Bacteriophages, commonly known as phages, are viruses that infect and replicate in bacterial cells. They are believed to be the most abundant organisms on the planet. One of the characteristic features of phages is that they are typically highly specific with regards to which hosts they can infect, often having species-level specificity or greater. This high specificity is based on selective binding of the virus ligand with the receptor at the bacterial surface (11). Given their abundance and specificity, phages are increasingly recognized as versatile tools with promising implications across multiple domains, including diagnostics, therapy for bacterial infections, environmental monitoring, and bacterial control in various settings (12,13).

The history and challenges of bacteriophage detection methods highlight a shift towards more efficient and versatile techniques. Traditional methods, such as the double agar layer (DAL) technique pioneered in the mid-20th century, remain foundational but are limited by their time-intensive and laborious nature, as well as their inability to scale for high-throughput applications (14). Emerging needs in clinical, environmental, and industrial settings, such as rapid assessment of phage interference in diagnostics and monitoring of phage therapies, demand faster and more adaptable detection methods (15). In response, modern approaches employ advanced technologies like PCR, qPCR, Raman spectroscopy, immunoassays, MALDI-TOF, and various optical and electrochemical methods. While these newer methods offer speed and potential automation, they often require sophisticated equipment and incur higher costs, posing barriers to widespread adoption in routine monitoring applications (12,15,16). Despite their advantages, achieving the sensitivity and precision comparable to the DAL method remains a challenge for many of these advanced techniques. Nonetheless, ongoing research and development aims to bridge this gap, aiming to deliver robust, sensitive, and cost-effective phage detection solutions suitable for diverse practical applications.

We previously found that *S*. Typhi bacteriophages were abundant in water sources from a typhoid-endemic region and were not detectable in communities without *S.* Typhi circulation. Our results demonstrated that using plaque assays to screen small water volumes is an effective method for detecting *S*. Typhi phages (9). Building on this work, we have now developed a scalable, colorimetric assay for environmental surveillance of typhoid. This new assay is both sensitive and rapid, enabling the detection of phages in just 1 mL of filtered water within 6 hours. Additionally, the test is cost-efficient, making it an ideal solution for low-resource settings.

## Methods

### Bacterial strains, Vi phages, and culture conditions

The laboratory strain *Salmonella* Typhi Ty2 was used for phage propagation. Additionally, we adapted our experimental conditions to use the attenuated *Salmonella enterica* serovar Typhi strain BRD948, which harbors deletions in the *aroA*, *aroC*, and *htrA* virulence-associated genes. This strain is attenuated, heavily encapsulated, and sensitive to all clinically relevant antibiotics, making it suitable for use in a biosafety level 2 containment environment (17). Bacteria were cultured overnight using Luria-Bertani (LB) broth Miller (BD Biosciences, USA), at 37 °C with aeration. When propagation experiments were performed with *S.* Typhi BRD948, LB media was supplemented with 40 mg L⁻¹ of l-phenylalanine and l-tryptophan, and 10 mg L⁻¹ of p-aminobenzoic acid and 2,3-dihydroxybenzoic acid (aro mix) to support the growth of this auxotrophic strain. Phages used as our experimental positive control were isolated from clinical samples and correspond to an *S*. Typhi Vi phage collection from the class *Caudoviricetes*, designated types II, III, IV, V, VI, VII. Vi phages were obtained from Cambridge University, Cambridge, United Kingdom. The original sources of these phages date from the 1930s through 1955 (18).

### Colorimetric assay

#### Experimental Procedure

A graphical representation of the phage colorimetric assay is exemplified in **Figure 1A**. In a sterile falcon tube, 1 mL of filtered water sample was combined with 900 µL of LB-broth. In a separate falcon tube, a positive control was prepared by adding 20 µL (10^10^ PFU/ml phage stock concentration) of Vi phage stock to 1900 µL of LB-broth. Negative control tube was prepared with 1 ml of field negative control filtrate and 900 µL of LB-broth; 100 µL of a fresh host culture (OD 600 = 0.2) was added to the sample, negative control and positive control tubes and gently mixed. An additional falcon tube containing 2 mL media was used to evaluate contamination. The enrichment culture and controls were incubated at 37°C for a minimum of 2 hours. Following incubation, the enrichment culture (‘high titer’) and controls were filtered through a 0.22 μm syringe filter to obtain a sterile filtrate suitable for testing.

**Figure 1.**
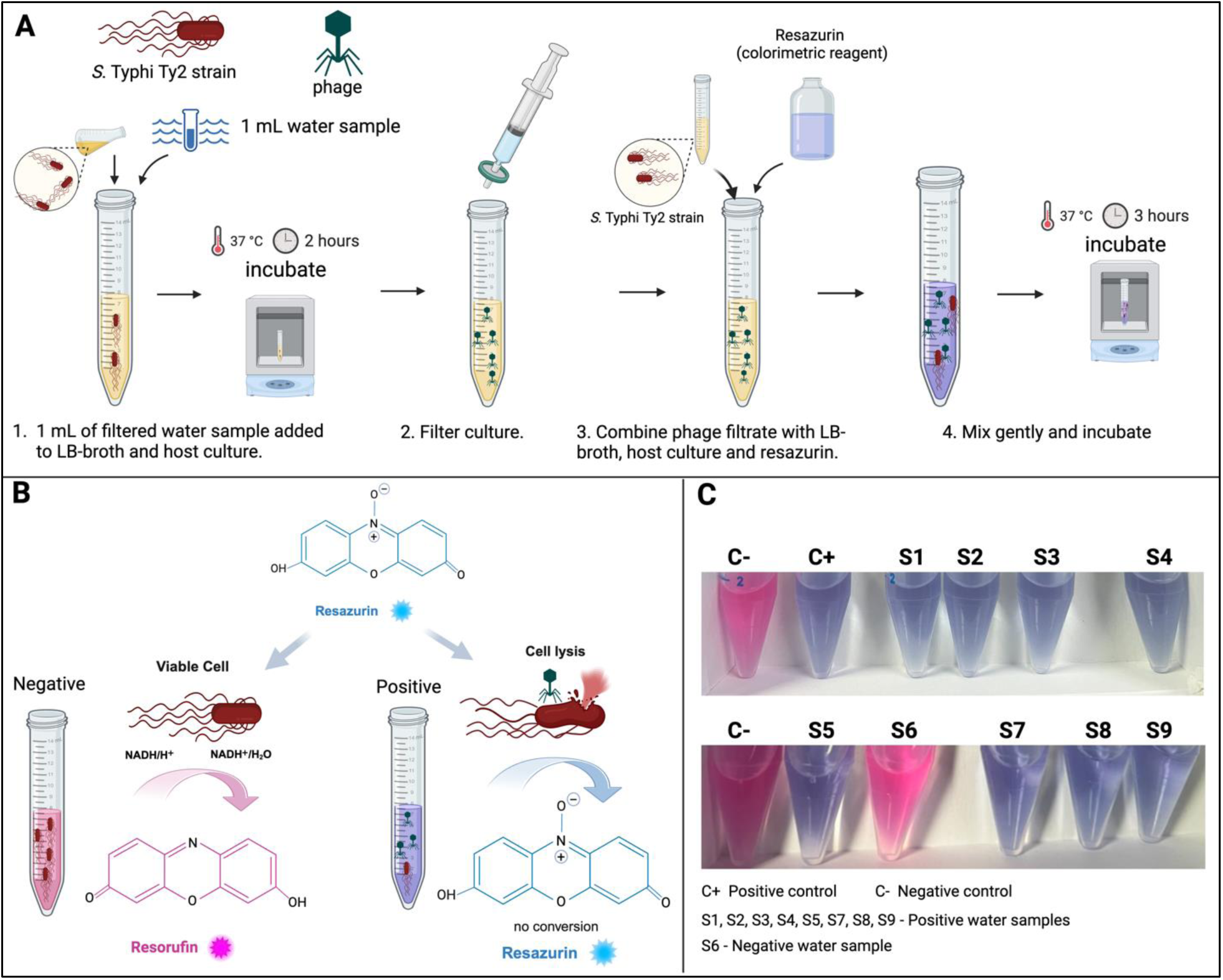
Phage colorimetric assay. **(A)** Schematic representation of the colorimetric assay procedure. **(B)** Illustration of the of the colorimetric reaction in the absence (pink color) or presence of *S*. Typhi phage (blue color). **(C)** Detection of *S*. Typhi in environmental water samples collected in Nepal. From left to right tubes **C-**, negative control (no sample addition), contains LB media, *S*. Typhi Ty2 strain and resazurin. **C+**, positive control (Vi phage control), contains LB media, *S*. Typhi Ty2 strain, Vi phage control and resazurin. **S1, S2, S3, S4, S5, S6, S7, S8, S** tubes contain LB media, *S*. Typhi Ty2 strain, different water samples and resazurin. Blue color indicates a positive result, and pink color indicates a negative result. Created with BioRender.com

In a new falcon tube, 100 µL of the enriched culture filtrate obtained in the previous step was combined with 1800 µL of LB-broth and, 100 µL of a fresh host culture (OD 600 = 0.2) and, 50 µL of AlamarBlue Cell Viability Assay Reagent (G-Biosciences Catalog #786-923), containing resazurin, and mixed gently. The culture was then incubated at 37°C for 3 hours. The assay is based on the resazurin cell viability method, which has been widely used for assessing cell activity and bacterial viability. Resazurin, a non-fluorescent blue dye, is reduced to resorufin, a pink and fluorescent compound, in the presence of metabolically active cells. The conversion is driven by cellular redox reactions, facilitated by NADPH dehydrogenase, which transfers electrons from NADPH to resazurin (Figure 1B).

The color change of the culture was monitored every hour during the incubation period. Positive results indicating phage presence were identified by a sustained blue coloration of the culture. Conversely, a change to pink color, reflecting bacterial growth uncontrolled by phage, indicated a negative result, reflecting the absence of lytic phage in the sample (Figure 1C). To enhance scalability and adaptability to resource limited settings, experiments were also performed with *S.* Typhi Ty2 lyophilized strain of which allowed us to avoid the need for overnight culture preparation of the strains. Lyophilization proved to be a valuable component, facilitating the use and reducing time requirements.

#### Colorimetric assay efficiency and limit of detection

To evaluate the efficiency and limit of detection of the colorimetric assay, we compared efficiency of phage colorimetric assay in tube format to a 96-well plate format and with phage detection by the standard double agar overlay. We performed the experiments using *S*. Typhi previously characterized Vi phage stocks (18). To obtain high concentration stocks of phages, we performed propagation assays using the double agar overlay method described below. Following overnight incubation clear zones were scraped and immersed in a 5ml SM buffer and kept for one hour at 4°C. The solution was then centrifuged at 4,000 rpm for 20 min and the supernatant was passed through a 0.22 µm syringe filter to obtain a sterile phage stock solution. Finally, the titer of stock solution was obtained by plating serial dilutions of the stock on a confluent lawn of the host bacteria and counting visible plaques to determine the approximate ratio of plaque-forming units per milliliter (PFU mL^-1^).

Overnight LB broth cultures of the host bacterium were used to perform all experiments. Phage stocks with a concentration of 2.8×10^10^ PFU/mL were first 10-fold serially diluted (10^-1^ to 10^-12^) in SM buffer. The colorimetric assay was performed as described above with all dilutions. The detection step of the colorimetric assay was also performed in a 96-well plate format. The plates were incubated at 37 °C in a BioTek Synergy H1 plate reader (Agilent Technologies, Vermont, USA) and fluorescence was recorded at regular intervals (every 30 minutes), with excitation set at 550 nm and emission set at 590 nm. The values obtained from the readings were expressed as relative fluorescence units (RFU). Samples, controls and blanks were assayed as triplicates. In a similar way, dilutions were also tested using agar overlay method. For these, 200 μL of *S*. Typhi overnight culture and 100 μL of the appropriate phage dilution were added to 4 mL of molten LB soft agar 0.7 % and poured into a petri dish containing solid LB agar. Each dilution was tested in three technical replicates. After an overnight incubation at 37 °C, plaques were counted. The PFU/mL from plaque assays were represented by the average of the three biological replicates.

In addition, we assessed assay stability by evaluating the optimal sample storage duration for phage detection. River water samples were collected in Kathmandu, Nepal and tested on the first day using both the colorimetric and agar overlay assays. The samples were then refrigerated at 4°C, and both tests were repeated every two days to assess how long the samples remained positive. This allowed us to compare the durability of assay detection in environmental samples and determine the efficiency and stability of the colorimetric assay over time.

#### Sampling sites

To evaluate the new assay in comparison with the double agar overlay method, we collected water samples from four countries with diverse geographic and epidemiological characteristics. We included sites where typhoid incidence rates are high or poorly documented, reflecting a spectrum of urbanization, sanitation infrastructure, and population densities.

*Nepal*. In July 2024, we conducted an environmental water sampling study in the Kathmandu Valley and Kavrepalanchok District of Nepal. Our focus was on five major rivers and one combined river segment, with samples taken from a total of 15 sites. Among the sampling sites 14 were located within the Kathmandu Valley and one was located in Kavrepalanchok. The site selection was guided by prior studies (9), emphasizing locations both upstream and downstream of river confluences. This sampling strategy was designed to comprise a variety of environments, from sparsely settled upstream areas to densely populated urban centers and downstream regions. Within the Kathmandu Valley, the majority of sampling sites (n = 10) were situated in or near Kathmandu Metropolitan City. The capital city of Nepal has a population of approximately 1,571,000 and a high population density of 17,103 people per km^2^. Unmanaged sewage, waste disposal, and lack of proper regulation in this highly dense area have led to their direct disposal into the river system in Kathmandu. Additionally, we sampled a river flowing through Banepa, a peri-urban city in the Kavrepalanchok District, with a population of 67,690 and a population density of 1,231 people per km^2^. Similar to Kathmandu, Banepa faces challenges with untreated waste and sewage entering its river systems. The estimated typhoid incidence rate in Nepal is relatively high, with previous studies reporting rates of 330 cases (230-480) per 100,000 people in Kathmandu and 268 cases (202-362) per 100,000 people in Kavrepalanchok (2).

*Côte d’Ivoire*. Environmental water sampling was conducted across various regions of Côte d’Ivoire, including both urban and rural areas. The primary urban center for sampling was Abidjan, the country’s largest city and economic hub. Sampling sites in Abidjan included diverse water sources, such as drainage ditches, open sewers, lagoons, streams, and areas where human activities (laundry, washing, etc.) were observed. Abidjan has a population of 6,321,017 and a population density of 2,934 people per km^2^. Wastewater in the city is often directly discharged into open drainage systems or natural water bodies due to insufficient infrastructure for sewage treatment. Additional samples were gathered from rural areas near Azaguie, Odoguie, and Alahin. In these rural areas, streams serve as vital water sources for the local population. Azaguie has a lower population density of 110 people per km^2^ and less infrastructure, which means people rely heavily on untreated water sources like streams. Here, contamination of water sources occurs primarily due to open defecation and lack of adequate sanitation facilities. In more rural areas like Azaguie, Odoguie, and Alahn, water samples were primarily taken from rivers and standing water. Similar contamination risks were observed due to poor sanitation practices. Typhoid fever incidence rates are not well-documented in Côte d’Ivoire. However, the Global Burden of Disease 2021 study estimated that Côte d’Ivoire has an annual incidence of 102 cases per 100,000 people (19).

*Niger*. In Niger, our environmental water sampling efforts concentrated on various locations, including urban centers like Niamey and rural areas such as Doguerawa, Galmi, and Magaria. In Niamey, the capital and largest city of Niger, water samples were collected from multiple sites including the River Niger, which flows through the city, and various drainage ditches. Niamey has a rapidly growing population, currently estimated at over 1 million residents with a population density of 1844 people per km^2^, which intensifies in the urban core. This density exacerbates the challenges of managing wastewater effectively. Despite some efforts to improve the sewage systems, many areas of Niamey still lack proper sewage infrastructure, leading to significant pollution of the river and drainage systems. In the rural areas of Doguerawa, Galmi, and Magaria, the sampling focused primarily on drinking water and drainage systems. These areas, characterized by low population density and minimal industrial activity, also face significant challenges related to water quality due to inadequate sanitation infrastructure. Similar to Côte d’Ivoire, the true number of typhoid cases in Niger is difficult to determine due to the absence of a coordinated epidemiological surveillance system. However, the occurrence of pathognomic ileal perforations throughout the country suggests that the incidence of typhoid is likely high (20).

*Brazil.* We conducted environmental water sampling from the primary watercourse in Campo Grande, the capital of Mato Grosso do Sul, Brazil. The city has a population of 898,100 and a population density of 111.11 people per km². The Anhanduí River, formed by the confluence of the Segredo and Prosa streams within the city’s urban center, is a canalized river that flows toward the southern region of the municipality, passing through 36 neighborhoods. We sampled five different sites along the Anhanduí River, each separated by approximately 3 km, both upstream and downstream from the river confluence. Additionally, we collected water samples from a park situated within the urban area, which features recreational zones. In this park, samples were taken from a canalized effluent that also discharges into the Anhanduí River. We also gathered sewage samples from the Sewage Treatment Plant (ETE) Los Angeles, which serves 90% of the city’s sewage collection and treatment needs. All collected sewage undergoes treatment at this facility before being discharged into the river. Typhoid fever is relatively uncommon in Brazil, with incidence rates having dramatically decreased since the 1980s.

However, cases still occur, particularly in the North and Northeast regions (21). In Mato Grosso do Sul, typhoid cases are very uncommon.

#### Sample collection and filtrate preparation

We collected 15 mL samples of water from the surface, against river current, in a sterile bottle, placed them into separate bags on ice, and transported them to the laboratory for further processing. A negative control sample was collected by pouring sterile distilled water into a bottle. Samples were passed through a 0.22 µm PES syringe filter (Sigma, USA) to obtain the filtrate, which was stored at 4°C until testing. At each sampling site, team members collected information on abiotic factors that may affect bacterial or viral survival, including solar UV exposure, presence of sewage pipes, open drain water carrying liquid and solid waste, and potential fecal exposure. Team members also recorded whether humans or animals were interacting with the water and the type of interaction that was observed.

#### Double agar overlay plaque assay

Phage screening was also performed by a double agar overlay method (14). For each sample, 1 ml of filtrate was added to 100 µl of overnight liquid bacterial culture and incubated at room temperature for 10 min to allow phage absorption. A positive control was prepared with 100 µl of Typhi-specific phage stock (10^4^ PFU/ml) and 100 µl of overnight liquid bacterial culture. A negative control tube was prepared with 1 ml of field negative control filtrate and 100 µl of overnight liquid bacterial culture. Molten Tryptic Soy Agar (0.7% agar) (3 mL) was added to the filtrate-bacteria mix and poured over solid hard LB agar base. Plates were incubated at 37℃ overnight and the presence of phages resulted in the production of visible plaques (zones of lysis) in a confluent lawn of the host bacterium.

#### Determination of phage host range

The lytic activity and specificity of phages was determined by screening each against different host bacterium strains using the standard double agar overlay method previously described. Serial dilutions of each phage stock were prepared and spotted on *S*. Typhi to obtain a concentration where plaques could be observed. Dilutions where plaques could be distinguished were used to spot phages on *S*. Typhi Ty2, *S*. Typhi Ty2 ΔVi (Vi capsule knockout mutant ΔtviB), *S*. Typhi ΔfliC (flagellate knockout mutant), *S*. Typhi CT18, *Salmonella* enterica serovar Paratyphi A (ATCC: 9150), *Salmonella* Paratyphi C (ATCC: 13428), *Salmonella* Choleraesuis (ATCC: 13312), *Salmonella* Enteritidis (ATCC: 13076), *Salmonella* Newport (ATCC: 6962), *Salmonella* Saintpaul (clinical isolate), *Salmonella* Typhimurium (ATCC: 700720), *Achromobacter xylosoxidans* (ATCC: 27063), *Acinetobacter baumannii* (ATCC: 19606), *Bordetella petrii* (ATCC:18323), *Citrobacter freundii* (ATCC: 8090), *Escherichia coli* (ATCC: 25922), *Enterobacter cloacae* (ATCC: 13047), *Klebsiella pneumoniae* (ATCC: 13883), *Morganella morganii* (ATCC: 25830), *Pseudomonas aeruginosa* (ATCC: 27853), *Proteus mirabilis* (ATCC: 29906), *Serratia marcescens* (ATCC: 13880), *Vibrio cholerae (*ATCC: 14035), and *Yersinia enterocolitica* (ATCC: 9610).

## Results

### Evaluation of the efficiency and limit of detection

To evaluate the efficiency of the colorimetric assay for detecting low concentrations of bacteriophages, we conducted a series of experiments with varying phage concentrations ranging from 0.028 to 2.8 x 10^10^ PFU/mL. The colorimetric assay demonstrated a detection limit of 28 PFU/mL (**Figure S1, A**), as evidenced by a noticeable blue color that was visible to the naked eye. At lower phage concentrations (2.8, 0.28, and 0.028 PFU/mL), the culture color closely resembled that of the negative control (no phage added), indicating that these concentrations were below the detectable threshold. We also assessed the impact of incubation time on the assay sensitivity. Cultures were observed at different time intervals (2:30, 3, and 3:30 hours post-incubation). The results indicated that the most distinct color differentiation between phage-positive and negative control cultures occurred at the 3-hour mark, leading to improved visual differentiation at this incubation time.

The assay was also performed in a 96-well plate format, where phage detection was assessed through the kinetics of fluorescence development due to resazurin reduction.

Fluorescence readings, measured as relative fluorescence units (RFU), were recorded at intervals until stable values were reached (**Figure S1, B**). For each concentration of phage, the Time of Detection (ToD) was calculated as the time required for the control (bacterial culture without phages) to reach the highest RFU value. At time 0, all phage concentrations, including the controls, exhibited similar baseline RFU values. After one hour of incubation, clear distinctions for fluorescence reading began to appear, especially at higher phage concentrations. Cultures with phage concentration of 2.8×10^9^ PFU/ml increased to 18,038 RFU, and at 2.8×10^8^ PFU/ml, the RFU reached 19,865, indicating the early stages of bacterial lysis. In contrast, lower concentrations (ranging from 2.8×10^3^ to 28 PFU/ml) remained relatively unchanged, with RFU values around 15,174 to 15,727. The ToD was identified as 2.5 hours, where most dilutions reached a plateau in RFU values. Cultures with phage concentrations between 2.8×10^9^ and 2.8×10^3^ PFU/ml showed substantial bacterial lysis, with RFU values ranging from 27,282 to 60,997, increasing as the phage concentration decreased. However, for dilutions of 2.8×10^2^ PFU/ml and lower, the RFU values either stabilized or reached the saturation point of 99,999. The limit of detection of the assay performed with 96-well plates format was established at 2.8×10^2^ PFU/ml, as dilutions equal to or greater than this value consistently reached the saturation point of 99,999 RFU, indicating minimal or no phage activity. In contrast, concentrations below this limit exhibited progressive increases in RFU, reflecting active phage-induced bacterial lysis.

The performance of the colorimetric assay was compared to the double agar overlay assay, which is considered the gold standard for phage detection. Results from both methods demonstrated 100% consistency in detecting *S.* Typhi-specific phages, particularly when the colorimetric assay was performed in a tube format. The plaque assay demonstrated the same limit of detection of 28 PFU/mL, with individual plaques still countable at this concentration (**Figure S1, C**). To further investigate the potential of the colorimetric assay as a scalable and low-cost method for environmental surveillance of typhoid fever, we compared its costs and resources with the phage plaque assay (the gold standard for bacteriophage detection) and PCR/qPCR, which is now the primary method for detecting *S.* Typhi in environmental samples. The results are summarized in Table 1.

**Table 1.**
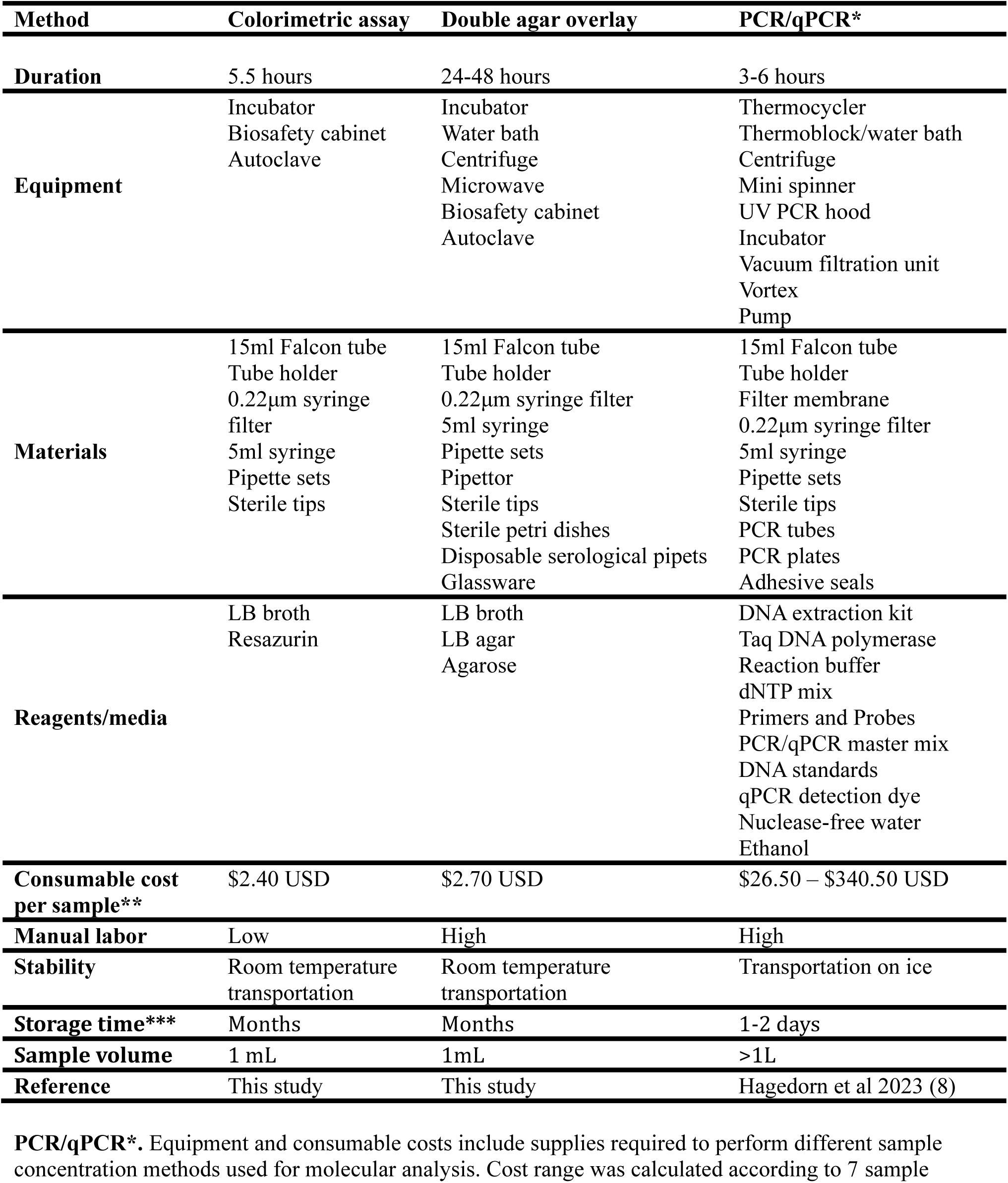
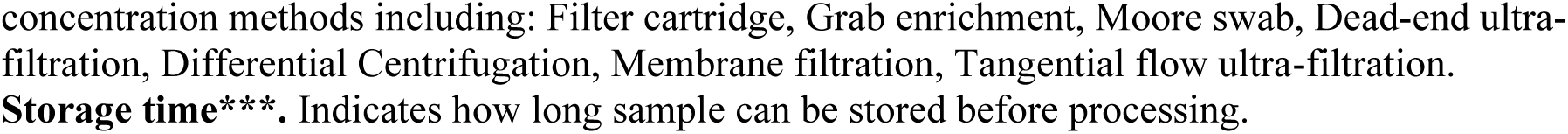
Costs and resource overview of typhoid environmental surveillance approaches.

| Method | Colorimetric assay | Double agar overlay | PCR/qPCR* |
| --- | --- | --- | --- |
| **Duration** | 5.5 hours | 24-48 hours | 3-6 hours |
| **Equipment** | Incubator<br>Biosafety cabinet<br>Autoclave | Incubator<br>Water bath<br>Centrifuge<br>Microwave<br>Biosafety cabinet<br>Autoclave | Thermocycler<br>Thermoblock/water bath<br>Centrifuge<br>Mini spinner<br>UV PCR hood<br>Incubator<br>Vacuum filtration unit<br>Vortex<br>Pump |
| **Materials** | 15ml Falcon tube<br>Tube holder<br>0.22µm syringe<br>filter<br>5ml syringe<br>Pipette sets<br>Sterile tips | 15ml Falcon tube<br>Tube holder<br>0.22µm syringe filter<br>5ml syringe<br>Pipette sets<br>Pipettor<br>Sterile tips<br>Sterile petri dishes<br>Disposable serological pipets<br>Glassware | 15ml Falcon tube<br>Tube holder<br>Filter membrane<br>0.22µm syringe filter<br>5ml syringe<br>Pipette sets<br>Sterile tips<br>PCR tubes<br>PCR plates<br>Adhesive seals |
| **Reagents/media** | LB broth<br>Resazurin | LB broth<br>LB agar<br>Agarose | DNA extraction kit<br>Taq DNA polymerase<br>Reaction buffer<br>dNTP mix<br>Primers and Probes<br>PCR/qPCR master mix<br>DNA standards<br>qPCR detection dye<br>Nuclease-free water<br>Ethanol |
| **Consumable cost per sample\*\*** | \$2.40 USD | \$2.70 USD | \$26.50 – \$340.50 USD |
| **Manual labor** | Low | High | High |
| **Stability** | Room temperature<br>transportation | Room temperature<br>transportation | Transportation on ice |
| **Storage time\*\*\*** | Months | Months | 1-2 days |
| **Sample volume** | 1 mL | 1mL | >1L |
| **Reference** | This study | This study | Hagedorn et al 2023 (8) |

**PCR/qPCR\*.** Equipment and consumable costs include supplies required to perform different sample concentration methods used for molecular analysis. Cost range was calculated according to 7 sample
concentration methods including: Filter cartridge, Grab enrichment, Moore swab, Dead-end ultra-filtration, Differential Centrifugation, Membrane filtration, Tangential flow ultra-filtration.
**Storage time\*\*\*.** Indicates how long sample can be stored before processing.

The colorimetric assay was the fastest method, requiring 5.5 hours to complete. This is considerably shorter than the 24 to 48 hours needed for the double agar overlay method, which is more time-intensive due to its incubation process. The PCR/qPCR method requires 3 to 6 hours, making it faster than the double agar overlay method but comparable to the colorimetric assay in terms of overall processing time. The colorimetric assay required minimal equipment, utilizing only an incubator, biosafety cabinet, and autoclave, which are standard items in most laboratory settings. In contrast, PCR/qPCR was the most equipment-intensive method, requiring sophisticated devices such as a thermocycler, thermoblock or water bath, centrifuge, UV PCR hood, and a vacuum filtration unit, along with specialized tools like a pump. These additional equipment requirements add significantly to both the cost and the operational complexity, making PCR/qPCR the most resource-demanding approach. The colorimetric assay proved to be the most cost-efficient, with a per-sample consumables cost of $2.40 USD. The double agar overlay method had a slightly higher cost of $2.70 USD per sample. PCR/qPCR, however, was significantly more expensive, with costs ranging from $26.50 to $340.50 USD per sample, depending on the method used for sample processing and DNA extraction.

In terms of manual labor, the colorimetric assay required low labor intensity, making it easier to implement and operate, especially in laboratories with limited resources. Both the double agar overlay method and PCR/qPCR were classified as high in terms of labor intensity, due to the multiple steps and complex procedures involved, which increase both the time and effort required for sample processing. For sample storage before processing, both the colorimetric assay and the double agar overlay method allowed for storage of up to 1 week. In contrast, PCR/qPCR samples had a much shorter storage window of 1-2 days, adding logistical challenges to its use, especially in field or low-resource environments.

#### Colorimetric detection of bacteriophages in environmental samples

A total of 182 environmental water samples were collected from Brazil, Côte d’Ivoire, Nepal and Niger (**Table 2**; **Figure 2**).

**Figure 2.**
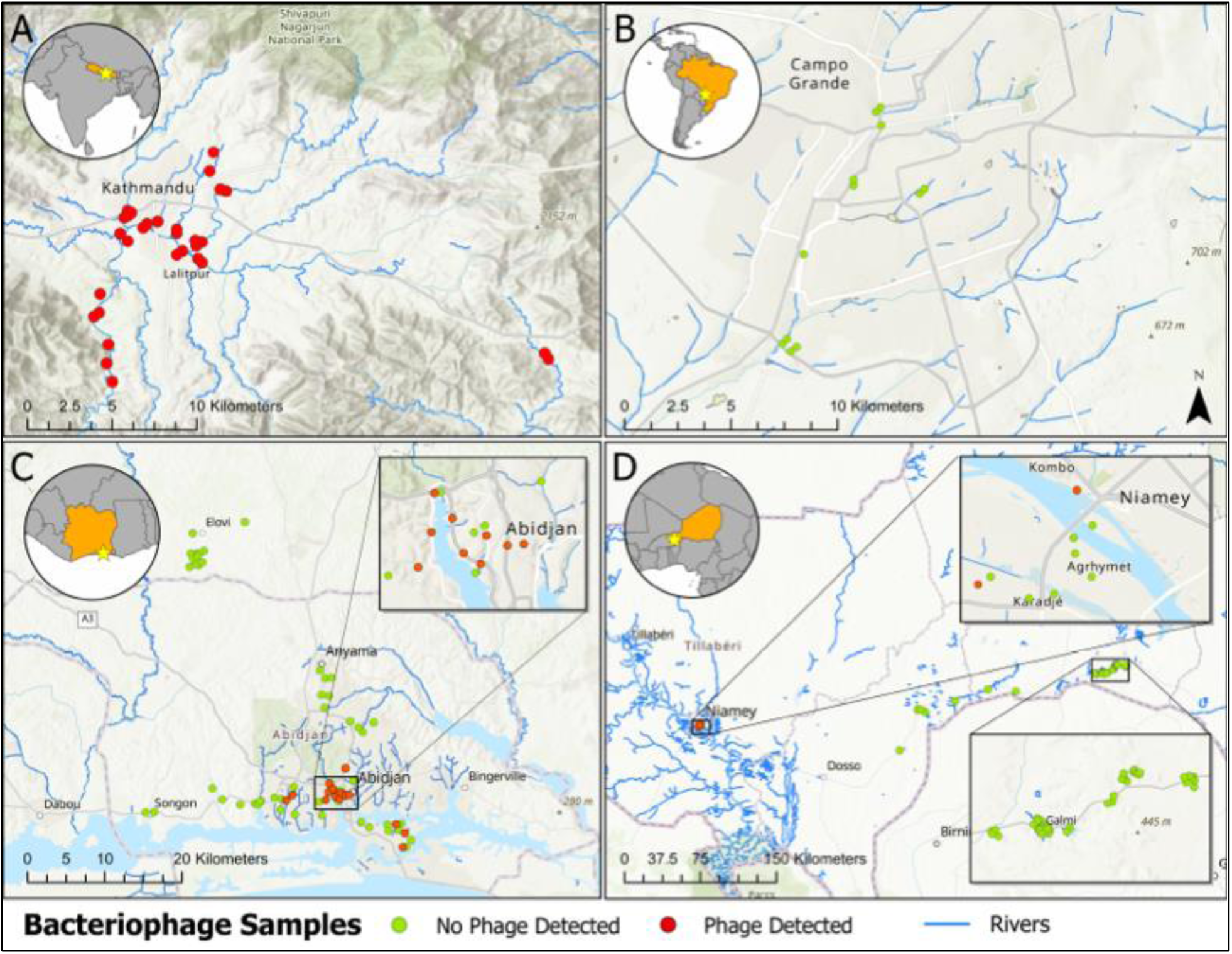
Water sampling location and detection of Salmonella Typhi phages from our study. (**A)** Kathmandu Valley and neighboring Kavrepalanchowk district in Nepal**. (B)** Campo Grande, Mato Grosso do Sul, Brazil. (**C**) Abidjan and neighboring cities in Côte d’Ivoire. (**D**) Niamey and the rural areas Doguerawa, Galmi, and Magaria in Niger.

**Table 2.**
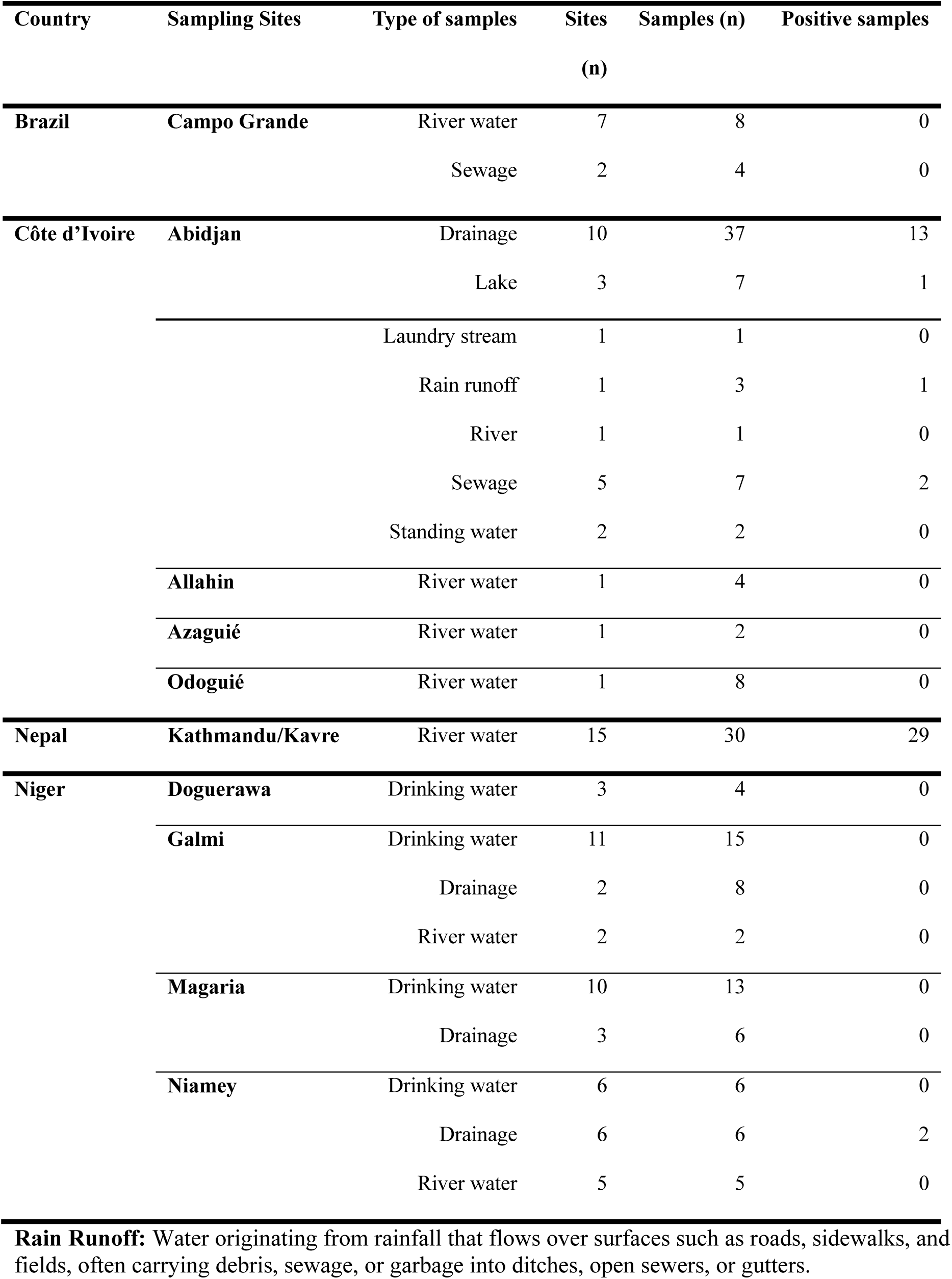
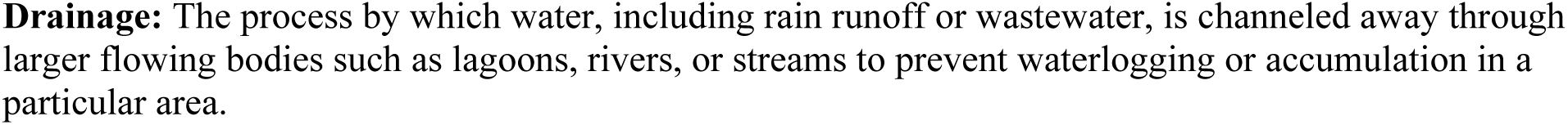
*S*. Typhi phage isolation from environmental samples collected in study sites.

| Country | Sampling Sites | Type of samples | Sites<br>(n) | Samples (n) | Positive samples |
| --- | --- | --- | --- | --- | --- |
| **Brazil** | **Campo Grande** | River water | 7 | 8 | 0 |
|  |  | Sewage | 2 | 4 | 0 |
| **Côte d'Ivoire** | **Abidjan** | Drainage | 10 | 37 | 13 |
|  |  | Lake | 3 | 7 | 1 |
|  |  | Laundry stream | 1 | 1 | 0 |
|  |  | Rain runoff | 1 | 3 | 1 |
|  |  | River | 1 | 1 | 0 |
|  |  | Sewage | 5 | 7 | 2 |
|  |  | Standing water | 2 | 2 | 0 |
|  | **Allahin** | River water | 1 | 4 | 0 |
|  | **Azaguié** | River water | 1 | 2 | 0 |
|  | **Odoguié** | River water | 1 | 8 | 0 |
| **Nepal** | **Kathmandu/Kavre** | River water | 15 | 30 | 29 |
| **Niger** | **Doguerawa** | Drinking water | 3 | 4 | 0 |
|  | **Galmi** | Drinking water | 11 | 15 | 0 |
|  |  | Drainage | 2 | 8 | 0 |
|  |  | River water | 2 | 2 | 0 |
|  | **Magaria** | Drinking water | 10 | 13 | 0 |
|  |  | Drainage | 3 | 6 | 0 |
|  | **Niamey** | Drinking water | 6 | 6 | 0 |
|  |  | Drainage | 6 | 6 | 2 |
|  |  | River water | 5 | 5 | 0 |

**Rain Runoff:** Water originating from rainfall that flows over surfaces such as roads, sidewalks, and fields, often carrying debris, sewage, or garbage into ditches, open sewers, or gutters.
**Drainage:** The process by which water, including rain runoff or wastewater, is channeled away through larger flowing bodies such as lagoons, rivers, or streams to prevent waterlogging or accumulation in a particular area.

Field validation of the phage colorimetric assay was performed in the Kathmandu Valley, Nepal. A total of 30 river water samples were collected from 15 sites, both within and downstream of densely populated areas. *S*. Typhi phages were detected in 29 of 30 samples (97%). In Abidjan, Côte d’Ivoire, a total of 58 samples were collected across several types of water samples, including drainage ditches (n = 37), sewage (n = 7), lake water (n = 7), and rain runoff (n = 3). Of these, *S*. Typhi phages were detected in 35.1% (13/37) of the drainage samples, 28.6% (2/7) of the sewage samples 14.3% (1/7) of the lake samples, and 33.3% (1/3) of the rain runoff samples. No *S*. Typhi phages were detected in river water (n = 14) samples from the rural regions, including Allahin, Azaguié, and Odoguié.

In Niger, a total of 65 samples were collected from drinking water (n = 38), drainage (n = 20), and river water (n = 7) across various urban and rural regions, including Doguerawa, Galmi, Magaria, and Niamey. *S*. Typhi phages were detected in 33.3% (2/6) drainage samples from Niamey, but none were found in any of the drinking water or river water samples across all regions. In Campo Grande, Brazil, 12 samples were collected from river water (n = 8) and sewage (n = 4) sites. No *S*. Typhi phages were detected in any of the samples.

#### Host range properties

To evaluate the host range and specificity of *S*. Typhi phages isolated, we tested all phages against a panel of relevant strains including additional *Salmonella enterica* serovars (Choleraesuis, Enteritidis, Newport, Paratyphi A, Paratyphi C, Saintpaul, and Typhimurium) and other bacteria such as *Achromobacter xylosoxidans, Acinetobacter baumannii*, *Bordetella petrii, Citrobacter freundii, Escherichia coli*, *Enterobacter cloacae*, *Klebsiella pneumoniae*, *Morganella morganii*, *Pseudomonas aeruginosa*, *Proteus mirabilis*, *Serratia marcescens*, *Vibrio cholerae* and *Yersinia enterocolitica*. Host ranges were determined by spotting phage solution onto bacterial lawns. The results show that most *S*. Typhi phages (n = 45) isolated in our study were only capable of infecting *S*. Typhi strains including a flagella knockout S. Typhi strain (ΔfliC). Although *S*. Typhi ΔfliC was susceptible to phage infection, no plaques were formed on the Ty2 isogenic Vi-negative *S*. Typhi strain (ΔtviB). Four phages isolated from samples collected in Nepal were able to infect both *S*. Typhi and *S*. Paratyphi A strains. None of the phages isolated were able to infect any other of the bacterial strains tested, including other *Salmonella enterica* serovars.

## Discussion

In this study, we developed a scalable, low-cost colorimetric assay for environmental surveillance of typhoid, focusing on the detection of *S.* Typhi phages in water sources.

Environmental water samples were collected from four countries—Brazil, Côte d’Ivoire, Nepal, and Niger—targeting regions with varying levels of urbanization, sanitation, and water management. Our results demonstrated a high prevalence of *S.* Typhi phages in water samples from densely populated urban areas, with the highest detection rates observed in Kathmandu, Nepal and Abidjan, Côte d’Ivoire. No *S.* Typhi phages were detected in river water from rural regions or drinking water samples across Niger and Brazil. The method has a detection range of 28 to 2.8 x 10^10^ PFU/mL and yields results in under six hours. These results support the potential of this assay as a low-cost, rapid method for identifying settings with *S.* Typhi circulating, which could inform the targeting of clinical and public health interventions including building medical laboratory capacity and vaccine introduction. Moreover, this pathogen-specific phage detection method offers a more sensitive and specific alternative to direct pathogen detection in environmental surveillance.

The findings of this study align with previous research linking poor sanitation and high population density with the presence of waterborne pathogens, including *S.* Typhi and its phages (6,7,9). The high detection rates of *S.* Typhi phages in urban areas like Kathmandu and Abidjan are likely due to untreated sewage entering open water systems, favoring the spread of *S.* Typhi and similar pathogens. This emphasizes the need to prioritize environmental surveillance in low-resource urban settings where overcrowding and inadequate waste management intensify contamination risks. Additionally, our findings are consistent with epidemiological data obtained from previous studies in Nepal, which demonstrated a high burden of typhoid fever in Kathmandu and Kavrepalanchok (2). These studies identified a positive association between population density and typhoid incidence, with higher rates observed in urban compared to peri-urban and rural communities (22,23). In contrast, the lower detection rates observed in Niger may reflect differences in sampling methods and environmental contexts. While urban areas in Nepal and Côte d’Ivoire featured dense populations and shared sewage networks that concentrated contamination, most samples from Niger were collected from water sources without clear fecal contamination. This likely led to lower positivity, despite epidemiological data suggesting a high typhoid burden in rural Niger. Future efforts should focus on identifying sewage sources to normalize sampling and enhance detection sensitivity.

The colorimetric assay demonstrated a sensitive limit of detection, consistent with the results obtained from double agar overlay method. When comparing the assay formats, the tube format proved to have superior detection sensitivity over the 96-well plate format. The limit of detection for the 96-well format was about ten-fold higher (280 vs 28 PFU/ml), likely due to the smaller sample volume, which could reduce the probability of capturing phages at lower concentrations. However, the 96-well format reduces personnel time when processing large numbers of samples, such that either approach could be useful depending on the setting and surveillance goals, weighing sensitivity and efficiency.

The colorimetric assay developed in this study offers a practical advantage for environmental surveillance, especially in settings where traditional detection methods may be unaffordable due to cost or complexity. Unlike the labor-intensive double agar overlay, which is time-consuming, our assay requires just 5.5 hours. Additionally, it avoids the need for expensive laboratory equipment, making it a more feasible alternative to PCR/qPCR, which, though highly sensitive, is costly and resource demanding, with per-sample costs ranging from $26.50 to $340.50 USD. These estimates exclude equipment expenses, which would make the differential costs even greater. The only major piece of equipment is an incubator, though we have previously shown that a low-cost, electricity-free incubator can be used (24). With a per-sample cost of just $2.40 USD, the most expensive component of our assay is the syringe filter.

However, there is significant potential to reduce costs further by exploring alternative filtration methods, as we have previously used in other applications (25). For instance, replacing syringe filtration with chloroform treatment reduces costs even further while maintaining performance. Such optimizations would make this assay even more accessible and scalable in resource-limited environments.

The phages isolated using the colorimetric assay demonstrated a high degree of specificity for *S.* Typhi strains, which is essential for establishing the assay as a reliable tool for environmental surveillance of typhoid fever. This specificity supports the potential use of these phages in targeted approaches for detecting *S.* Typhi in various environmental samples. Phages isolated in our study did not infect a Vi-capsule deletion strain, indicating the necessity of Vi capsule expression for phage infectivity, consistent with previous findings (18,26). Additionally we tested the phages isolated in the assay against other bacteria that were previously reported to express the Vi antigen (*A. xylosoxidans*, *B. petrii, C. freundii* and *S.* Paratyphi C). The phages were unable to infect these bacteria, demonstrating the assay’s high specificity for *S.* Typhi and its adaptability in distinguishing target pathogens within complex microbial environments.

Interestingly, four phages demonstrated the ability to infect both *S.* Typhi and *S*. Paratyphi A, despite the absence of the Vi capsule in *S*. Paratyphi A, suggesting that these phages may use alternative or additional receptors to mediate infection. In settings where *S.* Paratyphi A circulates (predominantly in Asia), adding the additional step of testing recovered phages against *S.* Paratyphi A would be important for determining whether *S.* Typhi-specific phages are circulating.

Despite the promising findings, several limitations should be acknowledged in this study. One key limitation is the absence of PCR/qPCR-based data to compare the accuracy and limit of detection between the colorimetric phage assay and more established molecular techniques. PCR is widely used for its sensitivity and specificity in detecting *S.* Typhi in environmental samples (5–7), and its inclusion in future studies would provide a valuable comparison for assessing the performance of the phage-based assay. However, we do note that given predator-prey dynamics of phages and bacteria, it is possible that at a sample level, *S.* Typhi and phage presence might diverge, even if phage presence indicates the circulation of *S.* Typhi within a community (27,28). Additionally, while the assay showed specificity for *S.* Typhi, further testing across a broader range of *S.* Typhi genotypes is needed to fully understand its range and possible cross-infectivity.

The detection limit of 28 PFU/mL, though sensitive, could also be further optimized to ensure its applicability in diverse environmental settings, especially where pathogen concentrations might be even lower. Furthermore, the presence of inhibitors, such as antibiotics or other antimicrobial substances in environmental samples, could interfere with the reliability of the assay.

Another limitation is that the assay does not measure the level of fecal contamination, which could provide crucial contextual information. Including coliform detection as an additional step would enhance the interpretability of the results, by enabling reporting of *S.* Typhi phage detection among samples that contain fecal contamination, providing a more consistent denominator across settings. Simple, low-cost methods already exist for coliform detection and enumeration in water samples (29). Additionally, future studies are required to correlate the frequency, levels, geographic distribution, and timing of phage detections with estimates of disease burden. Such correlations would provide important insights into the relationship between environmental phage presence and actual cases of typhoid fever, enhancing the assay’s value for public health surveillance. Finally, more detailed structural analyses of phage receptors and host interactions are required to clarify the mechanisms underlying the broader host range observed in some phages. Despite these limitations, these findings represent an important step forward to leverage phage detection as a proxy for pathogen detection in situations where direct pathogen detection is problematic.

In conclusion, the colorimetric assay offers a rapid, cost-effective, and scalable method for the environmental surveillance of *S.* Typhi bacteriophages. Its implementation in low-resource settings could provide an additional source of data to help guide typhoid control measures. Future studies should evaluate phage abundance alongside molecular-based environmental surveillance and clinical incidence studies. This approach may have the potential to reduce costs of environmental surveillance for *S.* Typhi and expand its accessibility in resource-constrained settings.

## Data Availability

All data produced in the present work are contained in the manuscript.

---

**Figure S1.**
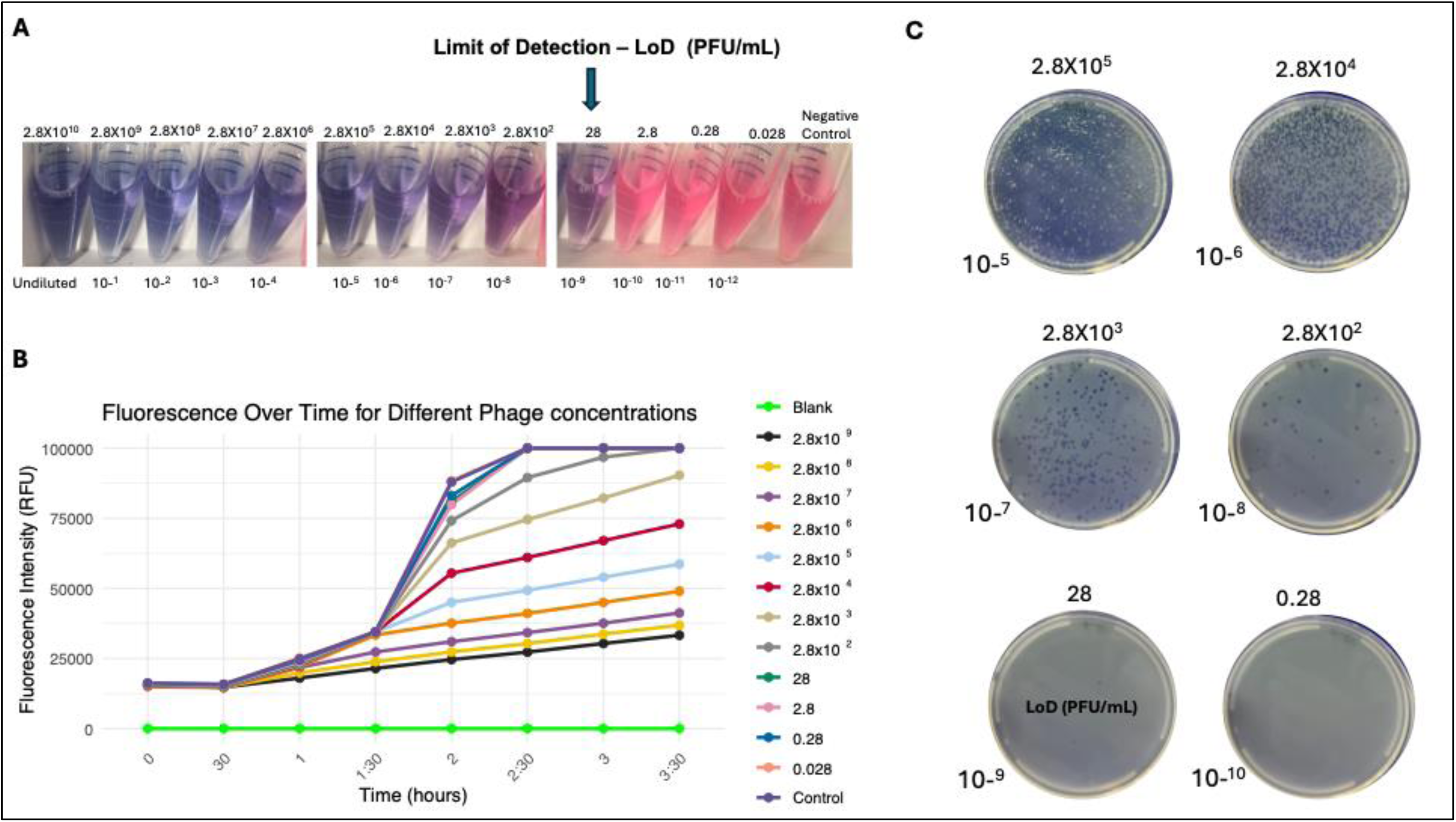
Evaluation of the efficiency and limit of detection of colorimetric assay. Several phage concentrations (0.028 to 2.8x10^-10^ PFU/ml) were tested and cultures contained LB media, *S*. Typhi Ty2 strain, Vi phage dilution and resazurin. Control, contains LB media, *S*. Typhi Ty2 strain and resazurin. (**A)** Colorimetric assay performed in the tube format. (**B**) Fluorescence kinetics in cultures of *S.* Typhi exposed to different Vi phage concentrations. Several phage concentrations (0.028 to 2.8x10^-9^ PFU/ml) were tested and cultures contained LB media, *S*. Typhi Ty2 strain, Vi phage dilution and resazurin. Control, contains LB media, *S*. Typhi Ty2 strain and resazurin. Blank, contains LB media. (**C**)

## References

1. GBD 2017 Typhoid and Paratyphoid Collaborators. The global burden of typhoid and paratyphoid fevers: a systematic analysis for the Global Burden of Disease Study 2017. Lancet Infect Dis. 2019 Apr;19(4):369–81.

2. Garrett DO, Longley AT, Aiemjoy K, Yousafzai MT, Hemlock C, Yu AT, et al. Incidence of typhoid and paratyphoid fever in Bangladesh, Nepal, and Pakistan: results of the Surveillance for Enteric Fever in Asia Project. Lancet Glob Health. 2022 Jul;10(7):e978–88.

3. Meiring JE, Khanam F, Basnyat B, Charles RC, Crump JA, Debellut F, et al. Typhoid fever. Nat Rev Dis Primer. 2023 Dec 14;9(1):71.

4. World Health Organization null. Typhoid vaccines: WHO position paper, March 2018 - Recommendations. Vaccine. 2019 Jan 7;37(2):214–6.

5. Andrews JR, Yu AT, Saha S, Shakya J, Aiemjoy K, Horng L, et al. Environmental Surveillance as a Tool for Identifying High-risk Settings for Typhoid Transmission. Clin Infect Dis Off Publ Infect Dis Soc Am. 2020 Jul 29;71(Suppl 2):S71–8.

6. LeBoa C, Shrestha S, Shakya J, Naga SR, Shrestha S, Shakya M, et al. Environmental sampling for typhoidal Salmonellas in household and surface waters in Nepal identifies potential transmission pathways. PLoS Negl Trop Dis. 2023 Oct;17(10):e0011341.

7. Uzzell CB, Abraham D, Rigby J, Troman CM, Nair S, Elviss N, et al. Environmental Surveillance for Salmonella Typhi and its Association With Typhoid Fever Incidence in India and Malawi. J Infect Dis. 2024 Apr 12;229(4):979–87.

8. Hagedorn B, Zhou NA, Fagnant-Sperati CS, Shirai JH, Gauld J, Wang Y, et al. Estimates of the cost to build a stand-alone environmental surveillance system for typhoid in low- and middle-income countries. PLOS Glob Public Health. 2023;3(1):e0001074.

9. Shrestha S, Da Silva KE, Shakya J, Yu AT, Katuwal N, Shrestha R, et al. Detection of Salmonella Typhi bacteriophages in surface waters as a scalable approach to environmental surveillance. PLoS Negl Trop Dis. 2024 Feb 8;18(2):e0011912.

10. Hooda Y, Islam S, Kabiraj R, Rahman H, Sarkar H, Silva KE da, et al. Old tools, new applications: Use of environmental bacteriophages for typhoid surveillance and evaluating vaccine impact. PLoS Negl Trop Dis. 2024 Feb 15;18(2):e0011822.

11. Jia HJ, Jia PP, Yin S, Bu LK, Yang G, Pei DS. Engineering bacteriophages for enhanced host range and efficacy: insights from bacteriophage-bacteria interactions. Front Microbiol [Internet]. 2023 May 31 [cited 2024 Sep 22];14. Available from: https://www.frontiersin.org/journals/microbiology/articles/10.3389/fmicb.2023.1172635/full

12. Elfadadny A, Ragab RF, Abou Shehata MA, Elfadadny MR, Farag A, Abd El-Aziz AH, et al. Exploring Bacteriophage Applications in Medicine and Beyond. Acta Microbiol Hell. 2024 Sep;69(3):167–79.

13. García-Cruz JC, Huelgas-Méndez D, Jiménez-Zúñiga JS, Rebollar-Juárez X, Hernández-Garnica M, Fernández-Presas AM, et al. Myriad applications of bacteriophages beyond phage therapy. PeerJ. 2023 Apr 21;11:e15272.

14. Ács N, Gambino M, Brøndsted L. Bacteriophage Enumeration and Detection Methods. Front Microbiol [Internet]. 2020 Oct 23 [cited 2024 Sep 22];11. Available from: https://www.frontiersin.org/journals/microbiology/articles/10.3389/fmicb.2020.594868/full

15. Paczesny J, Richter Ł, Hołyst R. Recent Progress in the Detection of Bacteria Using Bacteriophages: A Review. Viruses. 2020 Aug 3;12(8):845.

16. Richter Ł, Janczuk-Richter M, Niedziółka-Jönsson J, Paczesny J, Hołyst R. Recent advances in bacteriophage-based methods for bacteria detection. Drug Discov Today. 2018 Feb 1;23(2):448–55.

17. Tacket CO, Sztein MB, Losonsky GA, Wasserman SS, Nataro JP, Edelman R, et al. Safety of live oral Salmonella typhi vaccine strains with deletions in htrA and aroC aroD and immune response in humans. Infect Immun. 1997 Feb;65(2):452–6.

18. Pickard D, Toribio AL, Petty NK, van Tonder A, Yu L, Goulding D, et al. A conserved acetyl esterase domain targets diverse bacteriophages to the Vi capsular receptor of Salmonella enterica serovar Typhi. J Bacteriol. 2010 Nov;192(21):5746–54.

19. Piovani D, Figlioli G, Nikolopoulos GK, Bonovas S. The global burden of enteric fever, 2017–2021: a systematic analysis from the global burden of disease study 2021. eClinicalMedicine. 2024 Oct 18;77:102883.

20. Adamou H, Maman Bachir A, Sanoussi Y, Shafer K, Sukri L, Hobbs L, et al. Surgical Complications of Typhoid Fever: First National Typhoid Conference in Niamey, Niger [version 1; peer review: awaiting peer review]. VeriXiv [Internet]. 2024;1(25). Available from: https://verixiv.org/articles/1-25/v1

21. Silva Viana IP, Paulo Vieira C, Lima Santos Rosario I, Brizack Monteiro N, Sousa Vieira IR, Conte-Junior CA, et al. Typhoid Fever and Non-typhoidal Salmonella Outbreaks: A Portrait of Regional Socioeconomic Inequalities in Brazil. Curr Microbiol. 2024 Jan 9;81(2):57.

22. Andrews JR, Vaidya K, Bern C, Tamrakar D, Wen S, Madhup S, et al. High Rates of Enteric Fever Diagnosis and Lower Burden of Culture-Confirmed Disease in Peri-urban and Rural Nepal. J Infect Dis. 2018 Nov 10;218(suppl_4):S214–21.

23. Tamrakar D, Vaidya K, Yu AT, Aiemjoy K, Naga SR, Cao Y, et al. Spatial Heterogeneity of Enteric Fever in 2 Diverse Communities in Nepal. Clin Infect Dis Off Publ Infect Dis Soc Am. 2020 Dec 1;71(Suppl 3):S205–13.

24. Andrews JR, Prajapati KG, Eypper E, Shrestha P, Shakya M, Pathak KR, et al. Evaluation of an electricity-free, culture-based approach for detecting typhoidal Salmonella bacteremia during enteric fever in a high burden, resource-limited setting. PLoS Negl Trop Dis. 2013;7(6):e2292.

25. Ephraim RKD, Duah E, Andrews JR, Bogoch II. Ultra-low-cost urine filtration for Schistosoma haematobium diagnosis: a proof-of-concept study. Am J Trop Med Hyg. 2014 Sep;91(3):544–6.

26. Pickard D, Thomson NR, Baker S, Wain J, Pardo M, Goulding D, et al. Molecular Characterization of the Salmonella enterica Serovar Typhi Vi-Typing Bacteriophage E1. J Bacteriol. 2008 Apr;190(7):2580–7.

27. Faruque SM, Naser IB, Islam MJ, Faruque ASG, Ghosh AN, Nair GB, et al. Seasonal epidemics of cholera inversely correlate with the prevalence of environmental cholera phages. Proc Natl Acad Sci U S A. 2005 Feb 1;102(5):1702–7.

28. Pasricha CL, de Monte AJ, Gupta SK. Seasonal Variations of Typhoid Bacteriophage in Natural Waters and in Man, in Calcutta during the Year 1930. Indian Med Gaz. 1931 Oct;66(10):549–50.

29. Nelson CL, Fox TL, Busta FF. Evaluation of Dry Medium Film (Petrifilm VRB) for Coliform Enumeration. J Food Prot. 1984 Jul;47(7):520–5.

